# Ultra fast oscillations in the human brain and their functional significance

**DOI:** 10.1101/2023.02.23.23285962

**Authors:** Milan Brázdil, Vojtěch Trávníček, Jonathan Curot, Martin Pail, Monika Służewska-Niedźwiedź, Robert Roman, Filip Plešinger, Emmanuel Barbeau, Michal Kucewicz, Petr Klimeš, Jan Cimbálník, Pavel Jurák, William C. Stacey, Gregory A. Worrell

## Abstract

Human brain cell assemblies fire electrical impulses up to very high frequencies, which limit is not yet known. Using advanced intracranial microEEG recordings in a cohort of epileptic patients, we newly identified short-lasting oscillations in frequencies between 2 and 8 kHz. These ultra fast oscillations were consistently and locally detected in epileptic hippocampi but were extremely rare in non-epileptic brain tissue. The discovered electrophysiological phenomena thus may reflect neuronal hyperexcitability.

---

Using direct electrical signal recordings from the human brain via intracranial electrodes, a wide range of frequencies spanning from ultra slow (<0.5 Hz) to very fast oscillations (500-2,000 Hz) have been repeatedly detected (Brazdil et al., 2017; Worrell et al., 2012). All these electrophysiological phenomena can be observed in both healthy and functionally impaired brain structures using standard clinical macroelectrodes (1 - 10 mm^2^) to record local field potentials that reflected the summated synaptic and action potential activity, commonly called intracranial EEG (Buzsáki et al., 2012). In parallel, some studies with clinical macroelectrodes suggest that the human brain can generate even higher frequencies (above 2 kHz) (Sakura et al., 2009; Usui et al., 2010; Cao et al., 2014). If this is true, then EEG micro recordings, which remove volume averaging, should be a more potent tool to probe for these ultra fast frequency activities, as shown previously (Worrell et al., 2008, Brazdil et al., 2017). This study aimed to analyze resting-state intracranial EEG data from intrahippocampal hybrid (macro/micro) depth electrodes in a cohort of patients with epilepsy using high sampling frequencies (25, 30, or 32 kHz). Here we focused on EEG activities in between 2 and 8 kHz. Hypothetical ultra fast electrical events within the human brain might bear distinct functional significance when compared to lower frequencies and possibly could substantially contribute to our better insight into the pathophysiology of epileptic disorder on the microscopic level.

Using a spectrogram based detector, we analyzed EEG data from 466 microcontacts (199 “shafts”, 132 “bundles”=microwires, and 135 tetrodes) and 434 macrocontacts of 52 hybrid depth electrodes and 31 macroelectrodes in total, recorded in fifteen subjects who were investigated in four independent epilepsy monitoring units. As expected, in both macro- and microcontacts of the depth EEG electrodes implanted into the mesiotemporal structures, and mainly into the epileptic hippocampus, we repeatedly detected high-frequency oscillations (HFOs) in the frequency range of ripples (80-250 Hz) and fast ripples (250-500 Hz). We also detected very high-frequency oscillations (VHFOs) in a range of very fast ripples (500-1,000 Hz) and ultra fast ripples (1-2 kHz)(Usui at al., 2015, Brazdil et al., 2017, Revajova et.al, 2023). In addition, short-lasting oscillations in frequencies between 2 and 8 kHz were occasionally detected in the microrecordings from all of the investigated subjects. Two distinct types of these ultra fast oscillations (UFOs) could be differentiated and both types were observed in recordings across four independent EEG labs (Fig. 1). Type I UFOs had a pattern of typical spindle-like ripples, they were not accompanied by synchronous lower frequency activities, and their mean amplitude was 244.03±564.0 μV. Type II UFOs started with an initial “high amplitude” sharp deflection followed by an oscillation rapidly decreasing in its voltage. Also, type II UFOs mostly occurred independent of lower frequency activities (ripples, fast ripples, very fast ripples, and ultra fast ripples) and the mean amplitude was 824.2±1107.5 μV. They were less frequent when compared to UFOs I (330 versus 2085, p<0.001). Both types were found in 12/15 patients, while the remaining 3 only had Type I UFOs detected. Notably, UFOs fluctuate in character, time and location: different frequencies and types can occur in the same channel and both independently or synchronously in neighboring channels (Fig. 1D). In the time domain, 14.29±24.34% of ultra fast oscillations overlapped with spikes, and more rarely they overlapped with ripples (6.30±8.80%), fast ripples (6.08±8.90%), and very fast ripples (6.28±7.47%).

**Figure 1.**
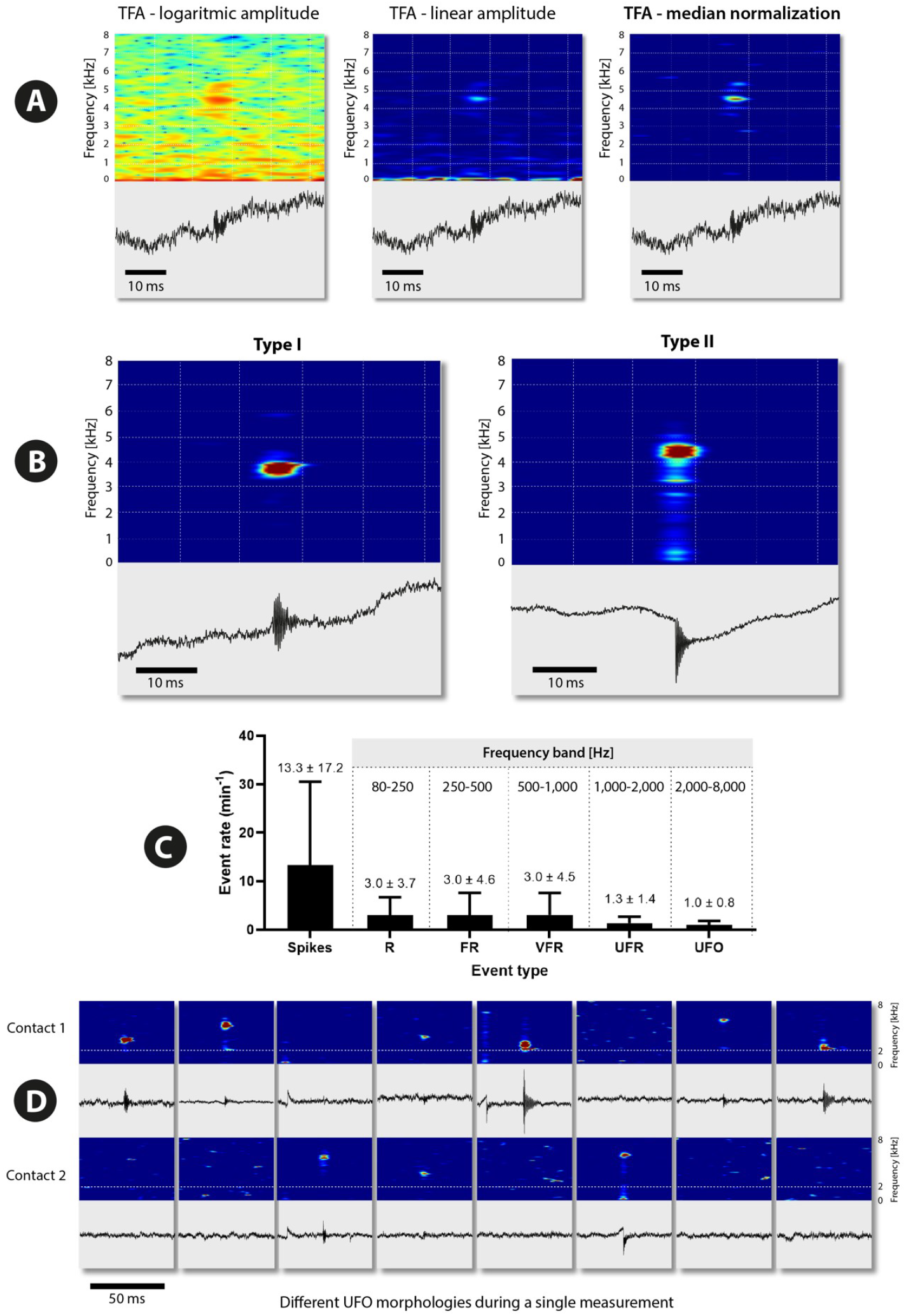
Visualization of raw signals with ultra fast oscillations in different spectrogram settings (median normalization used in all the time frequency maps in the paper) (A), examples of types I and II (time frequency maps and correlating raw EEG data) (B), relative rates of spikes, ripples, fast ripples, very fast ripples, ultra fast ripples and UFOs across all multi subject microrecordings (C), and example of mutual temporal relationships of distinct oscillations in two selected adjacent and simultaneous microrecording channels (D)

Importantly, when restricting the analysis to standard (macro) depth electrodes that had high enough sampling rate, HFOs over 2 kHz were not detectable except in a single contact in one of the three available patients. We verified that these were UFOs by comparing them with neighboring microelectrodes. However, the rate of UFOs on the macro contact was nearly 50 times smaller than on adjacent micro contacts (0.08/min versus 3.8/min).

In our study, ultra fast oscillations were consistently and locally detected in microrecordings from obviously epileptic hippocampi (in all five available intrahippocampal recordings), and once from a neocortex microelectrode adjacent to the epileptic hippocampus. A maximum of microoscillations was observed here in the frequency range of 2-5 kHz, but in four subjects they reached up to 7, and 8 kHz, respectively (Figs 2,3).

**Figure 2.**
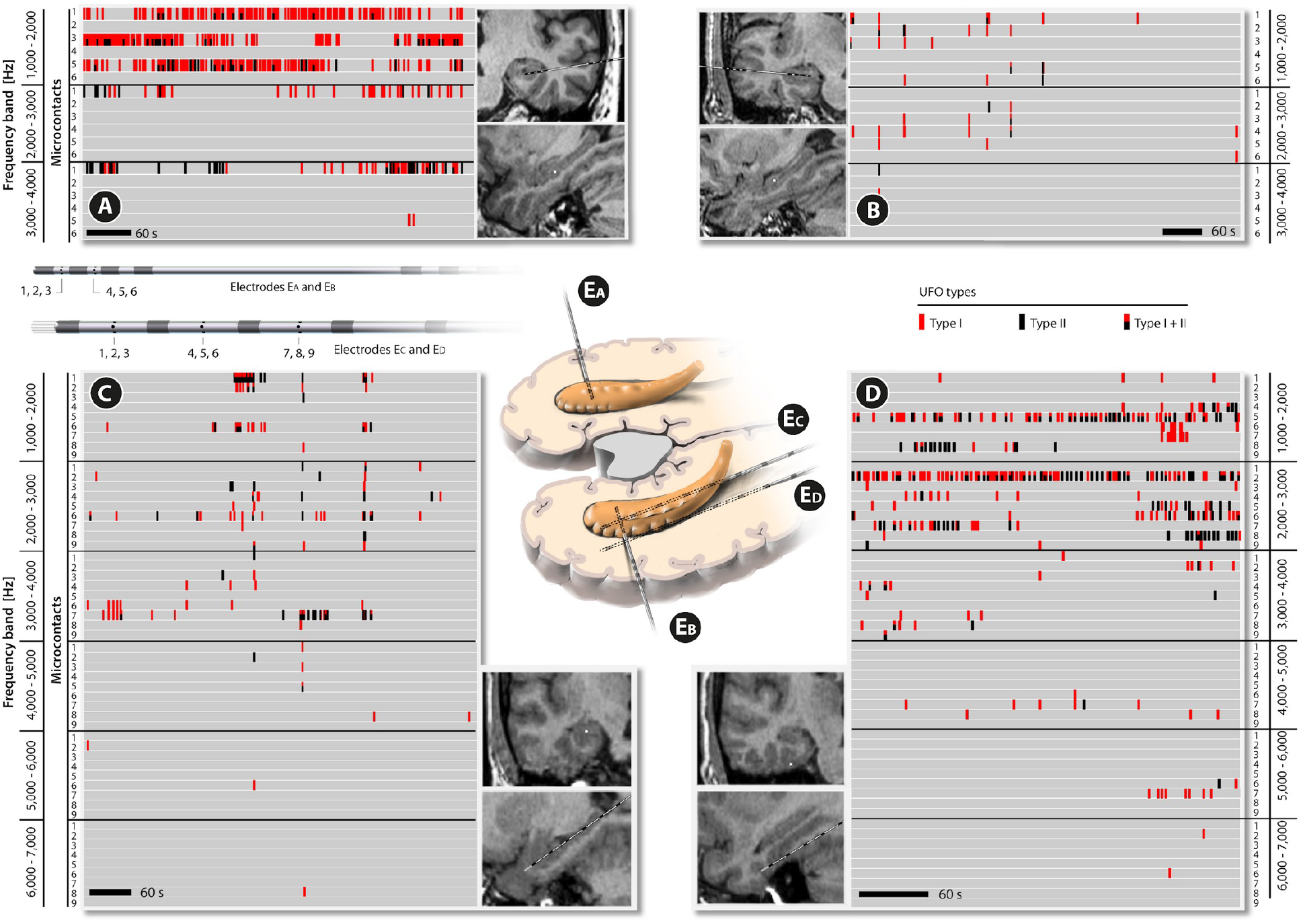
Distribution of UFR (1-2 kHz) and UFOs (2-8 kHz) across microcontact recordings in four investigated patients (A-D). For each subject one electrode data are shown only. A, C - epileptic hippocampus, B - non-epileptic hippocampus, and D - epileptic entorhinal cortex. Event detections for each microcontact in time are given in color strips reflecting the event type (I, II or both). Each strip is representing 1 sec time interval with at least one (or more) microoscillation.

**Figure 3.**
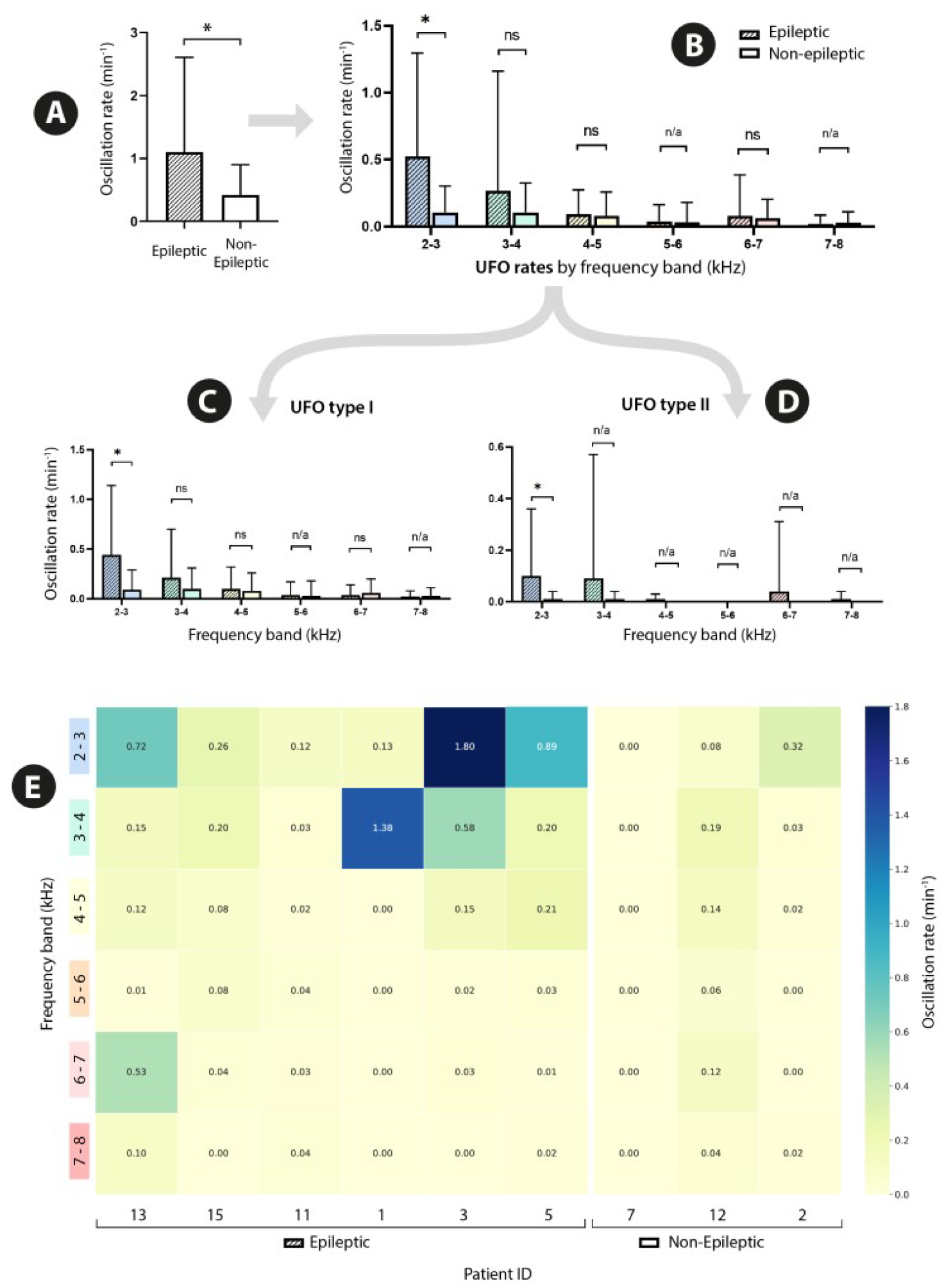
UFO rates in epileptic versus nonepileptic mesiotemporal regions, across 9 subjects. Broad frequency range 2-8 kHz (A), and more selective comparisons of epileptic versus nonepileptic ultra fast oscillations in investigated subjects (B). Rates of UFOs separately for type I and type II (C,D). Mean UFO rates (whiskers represent standard deviation), by frequencies across all microchannels within epileptic versus nonepileptic mesiotemporal regions (E). *:p<0.05, ns: p>0.05, n/a: not enough data for fitting the model.

The UFO rates varied inter-individually, across time, and recording microcontacts, with a maximum rate of 1.2-15.9/min for most active channels within six definitely epileptic structures (successfully operated). The ultra fast oscillations were present both widely across several recording microcontacts (generally in lower frequency ranges, 2-3 kHz) as well as more locally, and repeatedly even in a single contact (in higher frequency ranges). Importantly, Type I and Type II of ultra fast oscillations did not represent the identical phenomenon, even if both of them predominantly affected similar channels (Fig. 2). Finally, parallel recordings from adjacent macrocontacts consistently revealed traditional HFOs and VHFOs.

When comparing rates of UFOs (in broad frequency range 2-8 kHz) within epileptic (N=6) versus non-epileptic (N=3) mesiotemporal areas (hippocampi and entorhinal cortex), a mixed-effects model with patient as a random factor revealed significantly higher UFO rates in epileptic tissue (p < 0.05; Fig. 3). A similar result was present in the 2–3 kHz sub-band (p < 0.05), while a similar—but statistically non-significant—decrease was observed in the 3–4 kHz range (estimate = 2.16 vs 0.45; p = 0.125). No convincing or only very rare ultra fast oscillations were detected in non-epileptic hippocampi (N=3).

Despite a prevailing assumption that ultra fast EEG activity has its limitation at around 1,000 Hz and faster frequencies might be nothing but “neuronal chatter” (Niedermeyer, 2005), several papers published in the last decades are telling us the opposite. A novel ultra fast electrophysiological component of somatosensory-evoked potentials (1,235 - 2,632 Hz) was directly recorded from the human brain by Sakura et al. (2009) and even with higher frequencies by Cao et al. (2014). Three other independent research groups provided convincing proof of non-event very high frequency oscillations up to 2,000 Hz in human epileptic brains (Usui et al., 2010, 2015; Brazdil et al., 2017, Guo et al., 2023, Revajova et al., 2023). By analogy, high frequency activities at 2,000 Hz were identified in *vivo* intrahippocampal electrophysiological recordings from a rat using a novel pseudo-wavelet approach, and these activities increased in prominence with epileptogenesis (Hsu et al., 2010). Finally, even a simple specialized electric organ of weakly electric fish *Apteronotus leptorhynchus* comprising neuron-like cells is able to produce physiological and intentional discharges up to 2.2 kHz (Albert and Crampton, 2005). In this special case, however, extreme neuronal firing seems to be in large part a result of the evolution of voltage-gated ion channel genes (Thompson et al.,2018). Our observation of ultra fast oscillations in the frequencies between 2 and 8 kHz, recorded using microelectrodes mostly from the human epileptic hippocampi and their surroundings, can be seen as the next breakthrough step moving the limits of electroencephalography and our understanding of electrical events in the human brain. Microelectrodes have much higher impedance than macroelectrodes and are highly prone to artifacts (Stacey et al., 2013). Thus, our recordings were obtained using reliable high impedance amplifiers, and analyses were done with great care, and it is therefore highly unlikely we faced some kind of “false” HFOs. The origin of SEEG data in four independent labs across the world and distinct EEG recording systems, together with intra-individually selected observations across all investigated subjects, characteristics of the events, and significant predilection of ultra fast oscillations to epileptic tissue, make the artificial provenance of UFOs hardly possible, at least for characteristic “spindle-like” UFOs type I.

Skepticism on the authentic origin of UFOs might be nevertheless augmented with our knowledge of an absolute inactive phase of neurons at about 2 msec. A single synchronized neuronal group can generate discharges with the upper limit at about 300 Hz. On the other hand, the network mechanisms underpinning fast ripple and faster oscillations could be explained by phase delay of different subpopulations of synchronized neurons, i.e. out-of-phase firing (Jiruska et al., 2017). Such desynchronized subpopulations can fire with frequencies up to 500 Hz or even higher. The computational model presented recently by Fink et al. (2015) showed a similar effect on cellular level with a simulated cluster of asynchronous, uncoupled network of pyramidal cells. When the inhibitory mechanisms in the neural tissue are impaired and the pyramidal cells become very active due to intense synaptic input, the multiunit out-of-phase firing can occur and produce very high frequencies, very likely far more than 1,000 Hz. When the pyramidal cells are desynchronized, the aggregate electrophysiological signal can result in ultra fast oscillations of any frequency which detection is then only limited with an advancement of recording devices. We used a computational model to simulate whether such high frequencies can be generated by physiological mechanisms. Using the methods described in Fink et al 2015, we simulated the output generated when a network discharge was generated by multiple clusters of synchronized pyramidal cells firing action potentials. Each individual cell in a cluster only fired once, while different clusters fired with a delay of 1/F (F = 1000, 2000, 5000, 10000). Under these conditions, in which the overall burst is comprised of a chain of individual synchronized clusters firing with a short delay, the model was able to generate clear oscillations up to 10,000 Hz. This mechanism of UFOs’ origin is further strongly supported by our recently published study, establishing a mathematical framework based on bifurcation theory and examining how a reduced neuronal coupling within an epileptic focus could lead to very high-frequency (VHFOs) and ultra-fast oscillations (UFOs) in depth EEG signals (Pribylova et al., 2024).

To conclude, in this paper, we describe for the first time ultra fast electric field oscillations far above 2 kHz in the human brain. These electrophysiological events can be reliably detected using advanced depth microEEG recordings and they likely mirror neuronal hyperexcitability and reflect the hypersynchronous firing of neurons and a collective oscillation created by summation of the crests of single unit neuronal firing activity. As such, their future studies may provide a deeper insight into the pathomechanisms of epileptogenic brain.

## Data Availability

All data produced in the present study are available upon reasonable request to the authors.

## Author contributions

Conceptualization, M.B., P.J., P.K., and J.C.; Data curation, V.T., J.C, M.P., M.S.N., R.R., and J.C.; Formal analysis, M.B., V.T., W.C.S., and M.P.; Funding acquisition, M.B., M.K., J.C., and E.B.; Investigation, R.R., M.P., and M.S.N., Methodology, P.K., J.C., V.T., P.J., W.C.S., and G.A.W., Project administration, M.P., J.C., and M.K.; Resources, M.B., and M.K.; Software, V.T.; Supervision, M.B., and G.A.W., Validation, P.K., P.J., J.C., W.C.S., and M.K.; Visualization, F.P., V.T.; Writing, M.B., V.T., W.C.S, and G.A.W.

## Online Methods

### Patients

Fifteen consecutive patients (8 females; 7 males) with drug resistant focal temporal or frontal lobe epilepsy (DRE: Commission on Classification and Terminology of ILAE, 1989) who underwent presurgical evaluation using intracranial stereo EEG recordings (stereo-electroencephalography, SEEG) with at least one hybrid depth electrode were consented and included in the study. All of them suffered from focal impaired awareness seizures and two of them additionally (plus) from focal to bilateral tonic-clonic seizures. Five patients were recorded in Mayo Clinic, Rochester, U.S.A., four patients in CHU Toulouse, France, three patients in Medical University Hospital Wroclaw, Poland and three patients were investigated in Brno Epilepsy Center, Czech Rep., E.U. Subjects ranged in age from 19 to 53 years (mean age 35 years, SD = 11). The main demographic and clinical characteristics of the involved subjects are shown in Table 1.

**Table 1.** Patients’ demographic and clinical characteristics.

| Article subject number | Institute | Sex | Age at Seizure Onset | Age at SEEG | Seizure Frequency/Monthly | MRI (signs of) | Implanted hybrid electrodes | Type and side of epilepsy | SOZ | Surgery/ histopathology | Outcome (Engel) (years) | Total microcontacts | Microcontacts in epileptic/ nonepileptic hippocampus |
| --- | --- | --- | --- | --- | --- | --- | --- | --- | --- | --- | --- | --- | --- |
| 1 | Brno | F | 16-20 | 46-50 | 2 | normal | RT | T / R | right hippocampus | right AMTR/gliosis | IA (5) | 6 | 6/epileptic |
| 2 | Brno | M | 21-15 | 26-30 | 30 plus | posttraumatic changes left lateral T | LT | T / L | left lateral T neocortex | left lateral T lesionectomy/ posttraumatic changes | ID (4) | 6 | 6/non-epileptic |
| 3 | Brno | M | 31-35 | 51-55 | 4 | left HS | LT | T / L | left hippocampus | left AMTR/HS | ID (3) | 6 | 6/epileptic |
| 4 | Rochester | F | 1-5 | 26-30 | 1 | bilat HS | RT, LT | T / bilat | bilateral; left and right hippocampus | NA |  | 34 | NA |
| 5 | Rochester | M | 16-20 | 36-40 | 8 | normal | LT | T / L | left hippocampus | left AMTR/gliosis | IA (7) | 34 | 16/epileptic |
| 6 | Rochester | M | 36-40 | 46-50 | 4 | normal | RT, LT | T / bilat | bilateral; left and right hippocampus | NA |  | 28 | NA |
| 7 | Rochester | F | 1-5 | 21-25 | 4 | left HS | LT, RT | T / L | left hippocampus | left AMTR/HS | IC (8) | 34 | 8/non-epileptic |
| 8 | Rochester | F | 16-20 | 31-35 | 6 | normal | RT | T / R + E / L | right hippocampus + left extratemporal | NA |  | 63 | NA |
| 9 | Toulouse | F | 6-9 | 16-20 | 15 | postoperative changes right temporal | RT, LT | T / R | right hippocampus, right temporal pole + right entorhinal cortex | right AMTR / HS | IB (5) | 35 | NA |
| 10 | Toulouse | M | 26-30 | 46-50 | 4-7 | normal | RT, LT | T / L | left anterior and medial temporal regions | left AMTR/HS+ gliosis | IA (2) | 36 | NA |
| 11 | Toulouse | F | 31-35 | 46-50 | 5-7 | right HS + posttraumatic changes right T | RT, LT | T / R | right hippocampus and amygdala | right AMTR/HS | IA (4) | 32 | 12/epileptic |
| 12 | Toulouse | M | 6-10 | 26-30 | 6 | subtle cortico-subcortical blurring within T poles bilat. | LT, RT | F / R | right anterior cingulate | Cortical resection/FCD | IA(2) | 32 | 16/non-epileptic |
| 13 | Wroclaw | F | 11-15 | 21-25 | 15 | cortico-subcortical blurring temporo-parietal right | RT | T / R | right amygdala + right entorhinal ctx | Cortical resection/FCD IIB | IB(1) | 26 | 8/epileptic |
| 14 | Wroclaw | F | 6-10 | 31-35 | 30 plus | cortico-subcortical blurring cingulate gyrus left | LT | F / L | left midcingulate ctx | Cortical resection/FCD IIB | IA(1) | 48 | NA |
| 15 | Wroclaw | M | 11-15 | 26-30 | 10-12 | cortico-subcortical | LT | T / L | left hippocampus and amygdala | AMTR/FCD IIA | IB(1) | 56 | 24/epileptic |

T – temporal; E – extratemporal; F – frontal; R – right; L – left; SOZ – seizure onset zone; HS – hippocampal sclerosis; ctx – cortex; AMTR – anteromesial temporal resection; FCD – focal cortical dysplasia.

For the comparison between epileptic and non-epileptic hippocampi, inclusion criteria were: (1) seizure-free outcome after surgery (Engel Class I) and (2) the presence of a microelectrode located either within the resected (epileptic) or the non-resected (non-epileptic) hippocampus.

All patients underwent a comprehensive presurgical evaluation, including a detailed history and neurological examination, brain magnetic resonance imaging (MRI), neuropsychological testing, and scalp and invasive video-EEG monitoring. The duration of clinical monitoring and the location and number of implanted electrodes were determined per clinical considerations. After invasive videoEEG monitoring, twelve patients underwent surgical intervention details of which are shown in Table 1.

The follow-up interval after epilepsy surgery was at least 1 year. The study was approved by the Ethics Committees of all participating institutions. All patients signed an informed consent form.

### SEEG recordings

**At Mayo Clinic**, custom hybrid depth electrode designs (AD-Tech Medical Instrument Corporation, Racine, WI) were used. These electrodes are based on standard four and eight contact clinical depth electrodes. The depth electrodes consist of a 1.3 mm diameter polyurethane shaft with Platinum/Iridium (Pt/Ir) clinical macroelectrode contacts; each clinical contact is 2.3 mm long with either a 5 or 10 mm center-to-center spacing (surface area 9.4 mm^2^). In addition to the standard clinical contacts, the 8 contact depth has 8-24 microcontacts (between 1-4 macrocontacts), and the 4 contact depth has 16 microcontacts, positioned radially along the shaft of the depth electrode. Each electrode has a bundle of 9 Pt/Ir microwires that protrude 1–3 mm from the shaft tip. The microwires are made of insulated Pt/Ir wire, 40 um in diameter.

The patients received one hybrid SEEG electrode in the temporal lobe bilaterally and in one patient subdural strips and other intracerebral electrodes were implanted for monitoring of the parietal and occipital lobe. The electrode position within the brain was verified using CT with electrodes in situ.

A clinical acquisition system (Neuralynx Inc., Bozeman MT) was used to acquire intracranial EEG at Mayo Clinic. The signal was acquired at 32 kHz sampling frequency. Macro EEG was subsequently filtered by Barlett-Hanning 1 kHz low pass filter and downsampled to 5 kHz sampling frequency. The stainless steal suture placed in the vertex region of the scalp midline between the international 10–20 Cz and Fz electrode positions was used as a common reference.

**In Brno Epilepsy Center**, aside standard depth multi-contact platinum electrodes with a diameter of 0.8mm, a macrocontact length of 2mm, and an intercontact distance of 1.5mm (Microdeep, DIXI Medical, BesanÇon, France or Depth Electrodes Range, ALCIS, BesanÇon, France), one custom hybrid depth electrode (2069-ECP-8C6-35T06-2G-26, ALCIS, BesanÇon, France) was used in each subject. This electrode has eight macrocontacts (each with surface area of 5 mm^2^) and six microcontacts positioned radially along the shaft (surface area of 1075 µm^2^) interspersed between 1. and 3. macrocontacts (Fig. 2). Each patient received 3 to 14 depth electrodes usually orthogonally in the temporal and facultatively in frontal lobes. Their position within the brain was verified using MRI and CT with electrodes in situ.

A 192-channel research EEG acquisition system (M&I; BrainScope, Prague, Czech Republic) was used for the recording of 30 minutes of awake resting interictal SEEG recordings with a sampling rate of 25 kHz and dynamic range of ±25 mV. The EEG acquisition unit was battery-powered to eliminate line noise and microelectrodes were connected to specialized high impedance amplifiers (>1 GΩ). We used standard epilepsy monitoring unit protocols, but emphasis was given to eliminating power sources of electromagnetic radiation and 50 Hz power grid influence. No special shielded environment was used. Ground electrode served as a reference for microcontacts, mean from all macrocontacts was used as a reference for macrocontacts.

**In CHU Toulouse**, the hybrid electrodes (DIXI Medical, France) consisted of a standard macroelectrode (diameter: 0.8 mm) equipped with two or three tetrodes that protruded up to 2 mm from the shaft between the first and second most distal macrocontacts. A micrometer screw was used to extend the tetrodes from the electrode shaft after implantation. The tetrodes protruded from the shaft at a 30° angle. Each tetrode was made of 4 tungsten wires, 20 μm in diameter. The theoretical surface of each microcontact was 6.28×10-4 mm2. Each tetrode was 70-80 μm in diameter. For hybrid electrodes equipped with two tetrodes, tetrodes were at a 180° angle in relation to each other. They also had nine 2 mm-long macrocontacts regularly spaced at intervals along the shaft according to the overall length of the electrode. Three models with 2 tetrodes were available in a total exploration length space between contacts (in mm) of: 33.2/1.9; 40.4/2.8; 50.8/4.1. The length of the electrodes was chosen according to the depth of the cerebral target. For hybrid electrodes equipped with three tetrodes, the tetrodes were at a 120° angle in relation to each other. They had six 2 mm-long macrocontacts, three distal and three lateral, all spaced 2 mm apart (3 models were available with exploration lengths of: 34, 42 and 50 mm). These three tetrodes cover a theoretical triangular surface of 2 mm2. The three-tetrode configuration was generally chosen when the electrode passed through white matter between the lateral and distal cerebral targets, where it is usually not useful to record EEG signal. Microelectrode recordings were obtained using a 64-channel Cerebus™ system (Blackrock Microsystems, Salt Lake City, UT, USA) with a 30 kHz sampling rate in two patients, and an Atlas™ system (Neuralynx, Bozeman, MT, USA) with a native sampling rate of 32 kHz, subsequently downsampled to 16 kHz, in two other patients.

At **Wroclaw Medical University Hospital**, custom hybrid depth electrode designs (AD-Tech Medical Instrument Corporation, Racine, WI) were used. These electrodes are based on standard four to eight contact clinical depth electrodes. The depth electrodes consist of a 1.3 mm diameter polyurethane shaft with Platinum/Iridium (Pt/Ir) clinical macro-electrode contacts; each clinical contact is 2.3 mm long with either a 5 or 10 mm center-to-center spacing (surface area 9.4 mm^2^). In addition to the standard clinical contacts, the 6 contact depth has 10 microcontacts (between macrocontacts 1-3) positioned radially along the shaft of the depth electrode. The 8-contact depth called the Benhke-Fried electrode has a bundle of 8 Pt/Ir microwires that protrude 1–3 mm from the tip of the electrode lead. The microwires are made of insulated Pt/Ir wire, 40 um in diameter. The patients were implanted with one or two of the 6-contact hybrid SEEG electrodes in the temporal or the frontal lobe and three to four of the 8-contact Benhke-Fried electrodes in the temporal cortex unilaterally in any one patient with other standard clinical depth electrodes implanted for monitoring of the other lobes. The electrode position within the brain was verified using CT with electrodes in situ. A clinical acquisition system (Neuralynx Inc., Bozeman MT) was used to acquire intracranial EEG at high sampling rates for research purposes. The micro-contact signal was acquired at 32 kHz sampling frequency whereas the micro-contact signals were downsampled to 5 kHz sampling frequency. A subgaleal strip of four macrocontacts was placed in the vertex region of the scalp midline between the international 10–20 Cz and Fz electrode positions for a common reference signal for the macro-contacts. Micro-contact signals were referenced locally to one of the micro-channels that showed no neuronal spiking or other neural activities.

EEGWave was used for interactive graphical analysis of large, multichannel data, custom Python scripts were employed for batch processing of data and event detection, and R software was used for statistical analysis.

## Detection methods

### Oscillations above 1 kHz

Detection was performed in a 30 sec sliding window. First, a spectrogram of the input signal was computed with the FFT window of 0.015 s with 97% overlap and detrending of the windowed signal. Every row of the spectrogram was divided by its median (normalized) in 1 sec sliding window (Fig 4B). Only frequencies 1-8 kHz were taken into account. Higher frequencies might include artifacts from the recording system and events from lower frequencies are out of the scope of this analysis. One dimensional detection signal was created by summing all rows of the filtered and frequency-bounded (1,000 – 8,000 Hz) spectrogram. 99 percentile multiplied by factor of 5 was used for peak detection on the one dimensional signal (Fig 4C). Peak areas bounded by its inflex points were marked as high frequency activity. For each putative oscillation detection two things were saved for further analysis:

**Figure 4:**
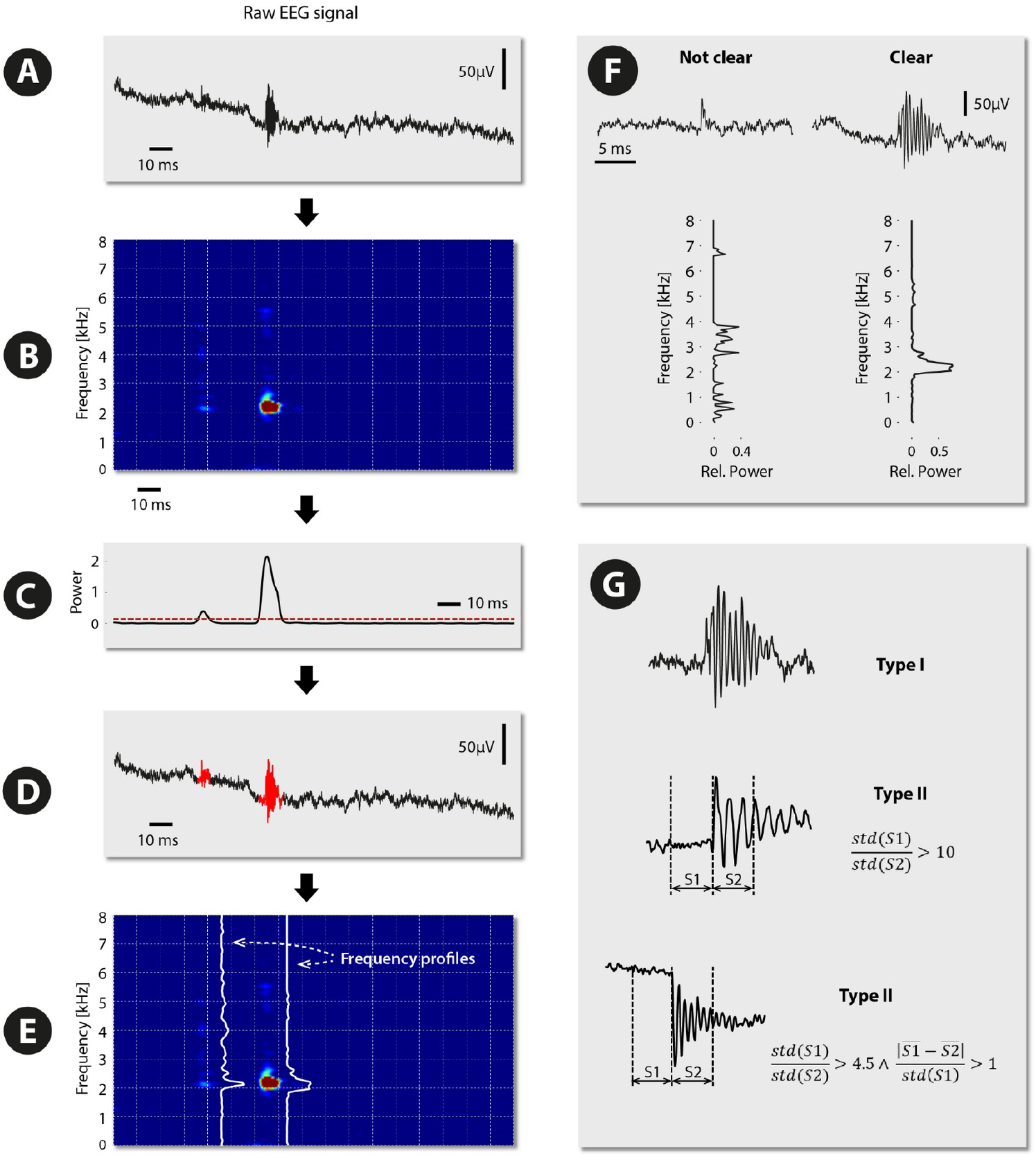
The figure introduces the UFO detection method. Primary detection of every high-frequency activity is depicted in the left panel. Part A) shows the raw signal. Its’ normalized spectrogram is in the B) part. Summing all rows of the spectrogram creates a 1-dimensional detection signal, which is thresholded (part C). D) part of the picture shows primary detections; its spectral profiles are shown in the E) part. The right panel shows filtering of primary oscillations. First, oscillations are assessed regarding their spectral profile as clear or not clear, which are removed from further analysis F). Other oscillations are sorted into two groups (part G).

1. Original raw signal of the putative detection
2. Spectral profile created by summing all columns of normalized spectrogram within time-bounded detection area (Fig 4E)

Every spectral profile of a detected high frequency event was subsequently classified either as biological or noisy. Event was marked as biological if the ratio of maximum to mean of the spectral profile was greater than 10. Other profiles were marked as noise because there was no dominant frequency and therefore could be considered as wide spread noise. Maximum to mean ratio served as the tuning parameter of the detector and was chosen heuristically. Lowering this number led to more detections, however their main frequency was less dominant. By setting this parameter to the value of 10, we chose 34.8±22.1% of primary detections, depending on the record (Fig 4F).

We observed that there were two major shapes of oscillations, Type I was fluently increasing and decreasing its amplitude and oscillates in one frequency band, which was represented by one circular or ellipsoidal area in spectrogram, while Type II oscillation started with sudden increase followed by slow decrease of amplitude often accompanied by shift of mean amplitude (see Fig. 4G bottom). This behavior led into one vertical strip in the spectrogram followed by high activity in one frequency area. To distinguish between these two shapes, the raw signal of every detection bounded by detection start and end was analyzed. The point where the absolute value of the derivative reached half of its maxima was marked as a True detection start. ¼ of detection length to the left from True detection start was marked as section 1 and ¼ of detection length to the right was marked as section 2. If the True start was not placed between ¼ and ¾ length of the detection it was automatically categorized as Type I. The nature of the detector (length of FFT window, using inflex points for detection start) ensures that Type II oscillations were always detected prior to their typical amplitude rise. Type II oscillation was defined by either of the following:

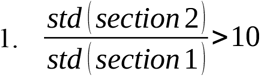

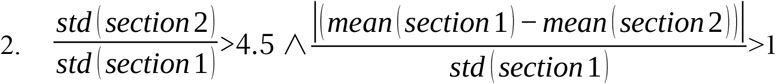

Equation 1. represents sudden change in standard deviation of amplitude within oscillation. So does equation 2., but one condition added, which represents a shift of means between two sections. Clear oscillations that did not fulfill any of these conditions were marked as Type I. Constants 10 and 4.5 can be considered as tuning parameters and were chosen heuristically. Using these constants we categorized apr. 35% of oscillations as Type II and 65% oscillations as Type I.

In some cases, the UFOs were detected simultaneously in more than three microcontacts at the same time. We declared these detections artifacts and omitted them from statistics because there is a risk that these oscillations are caused by electrode movement. 42% of all detections were declared artifacts.

To statistically compare the absolute count of Type I and Type II oscillation, we used non-parametric paired Wilcoxon signed-rank test, where oscillation count on each patient channel served as one sample.

### Ripples, fast ripples and very fast ripples

Oscillations in ripple (80-250 Hz), fast ripple (250-500 Hz) and very fast ripple (500-1,000 Hz) bands were detected using Line-length detector (Gardner et al., 2007). For every oscillation type, signal was first processed by 3rd order band-pass filter in respective band, detection was performed by Line-length detector with following parameters: the statistical window of 10 sec, 6 standard deviations was set as the detection threshold, window size was calculated as fs/50 samples, where fs stands for sampling frequency, window overlap was set to 25% of the sliding window length

### Spikes

Barkmeier detector (Barkmeier et al., 2012) was used for spike detection with following parameters: scale factor 70, threshold coefficient of 4 standard deviations, left and right slope ratios of 700, left and right spike duration of 0.1 second.

### Statistical analyses

All statistical analyses were performed using R software version 4.2.1. The significance threshold was set to α = 0.05 for all tests.

To compare the number of oscillations between epileptic and non-epileptic hippocampi, we employed a generalized linear mixed model (GLMM) implemented in the function glmmTMB() from the glmmTMB library (https://www.rdocumentation.org/packages/glmmTMB). The model accounted for repeated measurements from multiple electrodes within each patient by including patient identity as a random effect. Because the oscillation counts exhibited overdispersion, the negative binomial distribution was used.

For each analysis, the dependent variable was the number of detected oscillations per electrode in a 10-minute resting segment. The fixed effect was the group factor (epileptic vs. non-epileptic tissue).

Separate GLMMs were fitted for:

1. All UFOs combined, representing the total number of ultra-fast oscillations (2–8 kHz) detected across all channels
2. Type I UFOs, characterized by smooth spindle-like morphology and a single-band oscillation pattern
3. Type II UFOs, characterized by an initial sharp deflection followed by a decaying oscillatory burst

Each model was evaluated for six 1 kHz-wide frequency bands (2–3, 3–4, 4–5, 5–6, 6–7, 7–8 kHz) and for the broad band 2–8 kHz. For each band, the model estimated coefficients, 95 % confidence intervals (CIs), and p-values for the group effect.

Data adequacy was verified prior to testing. Bands with insufficient observations (typically > 5 kHz for Type II) were described qualitatively rather than modeled. Descriptive statistics were derived from model-based estimates and summarized as means with 95 % CIs.

The analytical design followed the same protocol across all datasets to ensure comparability. The limited sample size (9 patients: 6 epileptic, 3 non-epileptic) was recognized as a limitation of inferential power, particularly at higher frequencies.

